# Disposable versus reusable gastroscope in observation and endoscopic mucosal resection performance: a non-inferiority trial

**DOI:** 10.64898/2026.05.26.26354082

**Authors:** Mingtong Wei, Chenghai Liang, Huaqiang Ruan, Guolin Liao, Peng Peng, Xin Li, Jun Zou, Shiquan Liu, Ge Cao, Xianhuan Yan, Mengbin Qin, Jiean Huang

**Affiliations:** the Second Affiliated Hospital of Guangxi Medical University; Daichuan Medical (Shenzhen) Co., Ltd.

**Keywords:** Non-inferiority trial, Disposable gastroscope, Reusable gastroscope, Endoscopic Mucosal Resection, Disinfection

## Abstract

**BACKGROUND & AIMS:** Conventional reusable endoscopes incur significant expenses in the form of purchase, maintenance, reprocessing, and disinfection. Reprocessing is frequently ineffective even following the use of high-level disinfectants (HLDs). Disposable gastroscopy might be a strategy to decrease infectious outbreaks associated with reusable endoscope. The aim of this study was to analyze and evaluate the performance, efficiency and safety in gastroscopy observation and subsequent potential EMR procedure via the disposable gastroscope in a clinical setting.

**METHODS:** Patients who required gastroscopies and met the criteria were recruited to this prospective, open-label, non-inferiority study. After obtaining the written informed content, the enrolled subjects selected themselves independently to the disposable group or reusable group. The primary measure was to evaluate the acceptable image quality and whether the disposable endoscope devices could meet the basic clinical demands with a noninferiority margin of -8%. The second measures were to analyze and evaluate the image conditions, accepted endoscopic maneuverability, efficiency and safety of observation and advanced potential EMR procedure. Appropriate statistical methods were conducted via PASS software and SAS 9.4. A two-tailed *P* value < 0.05 was considered statistically significant.

**RESULTS:** A total of 90 individuals (the number of those in disposable group and reusable group was both 45) were recruited to this study. The success rate of acceptable image quality via photographing iconic anatomical sites between two groups was 100.0% (45/45, 95% confidence interval (CI): 0.9213,1.0000) and the lower limit of the 95%CI (-7.8654%, 7.8654%) was larger than the noninferiority margin of -8% (Newcombe-Wilson score method). Significant differences were showed in the measures of image conditions (image acquisition, image quality, brightness, contrast and sharpness) and accepted endoscopic maneuverability (endoscopy body rigidity). No significant differences were observed in the field of knob operation, sharp angle adaptability, and the auxiliary features including air supply, water supply and suction. In terms of efficiency, the total operating time, insertion time and withdrawal time were longer in the disposable group. The En-bloc resection rate of those observed polyps and required to EMR procedure due to relatively larger diameter (5mm-15mm) was the same 100% in both groups (26/26 vs 23/23, 95%CI: 0.8713,1.0000). Nevertheless, the procedure time of EMR for each polyp was significantly longer in the disposable group. This study showed no intraoperative bleeding, delayed bleeding, perforation or other study-related adverse events among 90 patients. No dramatic fluctuations in vital signs were showed in perioperative period.

**CONCLUSIONS:** In consideration of the efficiency, efficacy and safety evaluation, the disposable gastroscopes might represent an alternative to conventional reusable gastroscopes in routine examination and endoscopic mucosal resection.

## Introduction

Diagnostic and therapeutic procedures for patients with gastrointestinal diseases typically involve gastrointestinal endoscopy. It was estimated that approximately 60 million gastrointestinal endoscopic procedures are performed in China every year. Moreover, endoscopes incur significant expenses in the form of purchase, maintenance, reprocessing, and disinfection.

A gastric polyp is an abnormal growth of tissue from the surface of gastric mucosal membrane observed via endoscope examination. Gastric polyps reveal diverse pathologies and fundic gland, hyperplastic and adenomatous polyps are the three most recognized subtypes, and differences in environment, ethnic, geographic, gross aspect and clinical associations are observed among them. Most often, the gastric polyps are asymptomatic, but abdominal pain, bloating, nausea, vomiting, gastric outlet obstruction and chronic melena as iron deficiency anemia could be identified in part of the subjects. Gastric adenomatous polyp could be considered as part of a sequence leading from dysplasia to carcinoma. Also, the probability of clinical noteworthy signs and neoplastic transformation increases as size increases. In view of neoplastic and malignant potential, a more conservative approach taken by some physicians suggests that all polyps greater than 5 mm. should be eliminated completely^1^.

Endoscopic Mucosal Resection (EMR) is a well-established therapeutic technique which can be performed alternatively to surgery for suitable non-invasive lesions. It has been considered as a safe, efficient and cost-effective invention^2^. Ordinarily, EMR is conducted by reusable gastroscope, which was reprocessed to clean and disinfect according to the guidelines, standards or recommendations to minimize iatrogenic transmission risk. Nevertheless, a large number of endoscopic inventions are conducted in China annually which is accompanied by exorbitant costs^3^. Evidence has mounted that those reusable flexible endoscopes reprocessing is frequently ineffective even following the use of high-level disinfectants (HLDs), and the integrated factors might include complex structures, improper cleaning and repair issues^4^. Endoscope-induced infection remains a challenge.

The Coronavirus disease 2019 (COVID-19) pandemic imposed a severe burden among the whole world, especially in endoscopy diagnosis and therapy issue, and the viral transmissions by droplet, fomites or airborne approach during the endoscopic procedures are also seemed to be contributors. In 2019, the US Food and Drug Administration (FDA) issued that the medical institutions and manufacturers shall convert the reusable duodenoscope into partial or complete disposable duodenoscope to decrease the risk of infection. Subsequently, six single-use duodenoscope models had been approved by FDA in 2020 and several related public studies suggested that the same performance was provided as reusable duodenoscopes^5,6^.

`EndoFresh`^®^ disposable electronic gastroscope–XZING-W200B (Huizhou Xzing Technology Co., Ltd., China) with an electronic endoscope image processor (XZING-S2, serial number: `S221060101, S221060104`) has been recently developed. And the U.S. FDA and Conformite Europeenne (CE) have already cleared this device for the examination/therapy of the upper gastrointestinal tract. A prospective randomized study suggested that the XZING-W200B gastroscope could provide the same examining performance as reusable endoscope in terms of maneuverability and safety^3^.

The aim of this study is to evaluate the performance, efficiency and safety in gastroscopy observation and subsequent potential EMR procedure via the disposable gastroscope.

## Methods

### 1. Popularity

A total of 90 individuals (calculated by PASS 15 Software according to the initially endpoint with a non-inferiority margin of -8%) who met the criteria and hospitalized in the Second affiliated Hospital of Guangxi Medical University between Oct 7, 2021 and June 15, 2022, were recruited to this study. The inclusion criteria were as follows: (1) 18 to 75 years of age and no gender limit; (2) willingness to participate and written informed consent was obtained; (3) if gastric polyp was founded occasionally and considered in need of EMR, further criteria would be as follows: the diameter of polyp was ≥5mm but ≤15mm. What needed to emphasize was that EMR procedure was not obligatory in neither disposable group or reusable group. The exclusion criteria were as follows: (1) a history of anesthetics allergies; (2) accompanied with other malignancies; (3) incomplete lesion lifting after local injection; (4) a history of upper gastrointestinal surgery; (5) other clinical trial participated within 6 months of screening and follow-up process; (6) pregnant or nursing women; (7) contraindications to gastroscopy: ➀severe cardiovascular or cerebrovascular disorders; ➁acute or severe respiratory disorders or in febrile state within two weeks; ➂ systemic hemorrhagic disorders or coagulation abnormalities with hemorrhagic tendencies; ➃unstable vital sign or in shock condition;➄obstruction or perforation of gastrointestinal tract; ➅ giant gastrointestinal diverticulum; ➆ thoracic-abdominal aortic aneurysm; ➇ mental or intellectual disorders; ➈ severe spinal malformation; or (8) considered prudently by the investigator as unsuitable for the trial.

This non-randomized controlled study was approved by the Ethics Committee of the Second affiliated Hospital of Guangxi Medical University on Sept. 22^nd^, 2021. After obtaining the written informed content, the enrolled subjects would select themselves independently to the disposable group or reusable group one day before the gastroscopy.

### 2. Devices

The systemic devices included: (1) the XZING-W200B disposable gastroscope (Huizhou Xzing Technology Co., Ltd., China) with a visual image processor (XZING-S2, serial number: S22003003, S22003004); The major specifications: ➀ Optimal System: 110 ° field of view, forward viewing and the depth of focus is 3-100 mm in normal mode without near focus; ➁ Insertion section: Distal end outer diameter and Insertion tube outer diameter are both 11mm. Working length is 1300mm. ➂ The range of angulation could be up to 180° and down/left/right to 160°. (2) the reusable gastroscopes (GIF-H290, GIF-HQ290, Olympus Medical Systems, Tokyo, Japan) with visual image processor (CV-290), and light source (CLV-290SL) and water pump (OFP-2). A search could be conducted to find the relevant specifications at http://olympusmedical.com.hk/products/gastroenterology/gastroscopy/index.html)

### 3. Procedures

After recruiting, each patient was arranged to fast (no food or water) for at least 6 hours. As all devices had been placed, intravenous fentanyl citrate and etomidate anesthesia inducted simultaneously. The well-trained and experienced endoscopists (deputy chief physician or above) would conduct the operation. The images of 10 anatomical landmarks: esophagus, dentate line, cardia, fundus, gastric body, gastric angle, antrum, pylorus, bulb and descending duodenum would be captured by routine insertion and withdrawal procedures. If any lesion was observed, its morphology, size and site were photographed, identified and recorded. After the routine observation, completion of necessary biopsy and the polyp(s) which met the criteria as mentioned was/were founded, EMR therapy started. Injection methylthionine chloride and normal saline was used to create a submucosal cushion, separating the superficial gastric polyp from the underlying muscle layer to allow complete resection via the electronic diathermy snare. Blood pressure, heart rate, saturation of pulse oxygen and any clinical symptom were monitored and recorded throughout the whole process.

### 4. Evaluation measures

#### Primary measure

##### Acceptable image quality

Evaluation process: The entire procedure was videoed and recorded. The images of 10 anatomical landmarks as well as abnormalities were captured accurately. One or more representative images in each site would be captured and selected carefully for the gastroscopy report. For the evaluation, a clear image of each of these sites was used. Two researchers independently evaluated the gastroscopic images of the subjects enrolled. Third-party researchers resolved any discrepancies.

#### Secondary measures

##### 4.1 Image Evaluation

Image conditions: (1) image acquisition: whether successfully acquire the target images were recorded. (2) image quality: definition and whether the nature of lesion could be identified immediately were evaluated. (3) brightness, contrast and sharpness: whether the nature of lesions and the cavities could be identified were evaluated.

Evaluation criteria for the measures as the above: Each item was rated as A (good), B (relatively good), or C (poor). Clinical application was considered qualified if both operability and image quality were rated A or B; otherwise, it would be unqualified.

##### 4.2 Accepted endoscopic maneuverability

###### 4.2.1 Flexibility: including body rigidity, knob maneuverability and sharp angle adaptability were identified

###### 4.2.2 Auxiliary features included air supply, water supply and suction

Evaluation criteria for the aforementioned measures: Each item was rated as A (good), B (relatively good), or C (poor). Clinical application was considered qualified if both operability and image quality were rated A or B; otherwise, it would be unqualified.

##### 4.3 Efficiency of observation and potential EMR procedure

To evaluate the efficiency of observation, the total operation time, the insertion and withdrawal time were noted. Polyp would be detected in part of the enrolled subjects and polyp(s) detection rate was defined as the proportion of subjects whose polyp(s) was/were removed during screening endoscopy^7^. Diminutive polyp (<5mm in diameter) consists of the majority of polyps found during endoscopy, which can be removed via biopsy forceps. As for those subjects whose polyp(s) was/were founded and measured ranging from 5mm to 15mm, EMR procedure would be required and particularly focused in this investigation. The quantity and the procedure time were recorded. Total effective rate between two groups was evaluated and the criteria was defined as: (1) Effective: En bloc resection of the polyp(s). The rim of normal tissue should be included and the muscularis propria membrane was exposure over the base; (2) Ineffective: Incomplete tumor resection.

##### 4.4 Safety measures

###### 4.4.1 In-operation stability

It was defined as the stability of the individual’s blood pressure and heart rate. Details were recorded to calculate the stability rate. The in-procedure stability was evaluated by determining what percentage of patients had a change of blood pressure or heart rate that was greater than 20% from baseline^8^. Comparing with diastolic blood pressure, systolic blood pressure was focused and recorded as normal, between 140 and 160 mmHg, > 160 mmHg, or in shock; heart rate was noted as normal, > 100 bpm, < 60 bpm, or arrhythmia. After anesthesia, each subject was positioned in supine in order to measure blood pressure and heart rate before, during (until the gastroscopic tip reached the middle part of gastric body), and 10 ± 5 minutes and 1 hour ± 15 minutes after the procedure and the percentage of individuals who had a blood pressure or heart rate that fluctuated by more than 20% would be calculated^3^.

###### 4.4.2 Incidence of intraoperative bleeding

The proportion of subjects who was in intraoperative bleeding among the experimental group or control group. The criteria: obvious bleeding within 10 seconds after the EMR procedure.

###### 4.3.3 Incidence of perforation

The proportion of perforation occurring during the potential EMR procedure in all subjects. Evaluation criteria: severe abdominal pain, peritoneal irritation, and the presence of free gas indicated by abdominal upright and decubitus radiography or abdominal CT were considered as perforation.

###### 4.3.4 Incidence of delayed bleeding

In surveillance of potential delayed bleeding, the follow-up process would be acquired within 1 month after the EMR procedure. Whether did they have the symptom as hematochezia during the follow-up period would be recorded in detail.

###### 4.3.5 Incidence of equipment malfunction/defect

Whether there was any malfunction/defect of the gastroscope and affiliated devices such as image interruption, blockage or leakage of water delivery and so on during the whole procedure would be recorded in detail immediately.

###### 4.3.6 Incidence of adverse events except above

Any severe adverse event due to the observation or therapy procedure such as respiratory depression, shock/hypotension, myocardial infarction, asphyxia, GI stricture, fistula or sinus formation and so on would be recorded in detail immediately.

#### 5. Emergency Management

As risks of any clinical trial exist objectively, all emergency measurements adequately prepared. Once the adverse event occurred, Blood Transfusion Department, Surgery Department and Intensive Care Unit would be on standby and authentic information would be recorded without reservation. Therefore, all the enrolled patients would get guaranteed in health.

#### 6. Statistical analysis

Continuous variables were expressed as the mean and standard deviation (s.d.) or the median and IQR. Counts and percentages were determined if appropriate. Measurement data were analyzed with the t-test or Wilcoxon rank sum test for intergroup comparisons. The categorical variables were analyzed by Pearson’s chi-square test or Fisher’s exact test. A two-tailed *P* value < 0.05 was considered statistically significant (SAS version 9.4).

## Results

### 1. General characteristics

This prospective, open-label, non-inferiority study enrolled 90 subjects. In terms of age and gender, there were no significant differences between two groups. No previous anesthesia allergy or gastrointestinal surgery history were founded. Part of them in disposable group (4/45) and reusable group (6/45) (*P*>0.05) had the medical history such as Hypertension, Diabetes Mellitus, but they were all well under control via standardized drug therapies, we considered it no significant impact on gastroscopy. The incidence rate of the relative symptoms like abdominal pain, bloating, nausea or vomiting indicated no obvious differences. **(Table 1)**

**Table 1.** Baseline demographics and features of enrolled patients.

|  |  | **Disposable**<br>**Group**<br>**(N=45)** | **Reusable**<br>**Group**<br>**(N=45)** | **P** |
| --- | --- | --- | --- | --- |
| **Age** |  | 49.80±13.81 | 46.33±15.37 | 0.263 |
| **Gender** |  |  |  | 0.833 |
|  | Male | 21 | 23 |  |
|  | Female | 24 | 22 |  |
| **Previous anesthesia allergy** |  |  |  | NS |
|  | Yes | 0 | 0 |  |
|  | No | 45 | 45 |  |
| **Gastrointestinal surgery history** |  |  |  | NS |
|  | Yes | 0 | 0 |  |
|  | No | 45 | 45 |  |
| **Medical history** |  |  |  | 0.739 |
|  | Yes | 4 | 6 |  |
|  | No | 41 | 39 |  |
| **Abdominal Pain** |  |  |  | 0.783 |
|  | Yes | 9 | 7 |  |
|  | No | 36 | 38 |  |
| **Bloating** |  |  |  | 0.590 |
|  | Yes | 7 | 10 |  |
|  | No | 38 | 35 |  |
| **Nausea or vomiting** |  |  |  | 0.737 |
|  | Yes | 6 | 4 |  |
|  | No | 39 | 41 |  |

### 2. Evaluation measures

#### (1) Primary measure

Acceptable image quality: The success rate of acceptable image quality via photographing iconic anatomical sites between two groups was 100.0% (45/45, 95% confidence interval (CI): 0.9213,1.0000). The between-group difference was 0, and the lower limit of the 95%CI (-7.8654%, 7.8654%) was larger than the noninferiority margin of -8% (Newcombe-Wilson score method), which was the non-inferiority threshold, suggesting that the image quality in the disposable group was non-inferior to that in the reusable group **(Table 2)**.

**Table 2.** Comparison of the success rate of photographing iconic anatomical sites in two groups.

| **Group** | **Success rate** | **Difference** | **Difference**<br>**95% CI#** | **Noninferiority**<br>**margin** |
| --- | --- | --- | --- | --- |
| Disposable (n =45) | 100% | 0 | (-7.8654%,<br>7.8654%) | -8% |
| Reusable (n = 45) | 100% |  |  |  |

#### (2) Secondary measures

##### 2.1 Image conditions

There were significant differences between the control group and the experimental group in the index of image acquisition, image quality, brightness, contrast and sharpness (*P*<0.05). All grades were evaluated as A (good) and B (relatively good), which was considered qualified for clinical application. No C (poor) grade was marked. **(Table 3)**

**Table 3.** Comparison of image conditions between disposable group and reusable group.

|  | Disposable<br>Group | Reusable<br>Group | *P* |
| --- | --- | --- | --- |
| **4.1 Image conditions** |  |  |  |
| **Image acquisition** |  |  |  |
| A: Good quality | 34 | 43 | 0.016 |
| B: Relatively good quality | 11 | 2 |  |
| **Image quality** |  |  |  |
| A: Good definition and the lesion could be identified immediately | 30 | 41 | 0.010 |
| B: Relatively good definition and the lesion could be identified with close observation | 15 | 4 |  |
| **Brightness, contrast and sharpness** |  |  |  |
| A: Good definition. The lesions and the cavities could be identified immediately | 33 | 43 | 0.009 |
| B: Relatively good definition. The lesions and the cavities could be identified with close observation | 12 | 2 |  |

##### 2.2 Accepted endoscopic maneuverability

The rates (rating: A or B) of the following index were 100% (45/45, 95%CI: 0.9213,1.0000) in both groups. The control group had a better rating in terms of endoscopy body rigidity (*P*<0.05) but no significant differences were observed in the field of knob operation, sharp angle adaptability, and the auxiliary features including air supply, water supply and suction. **(Table 4)**

**Table 4.** The maneuverability Evaluation in the two groups.

|  | Disposable<br>Group | Reusable<br>Group | *P* |
| --- | --- | --- | --- |
| **4.2.1 Flexibility** |  |  |  |
| **Body rigidity** |  |  |  |
| A: Moderate rigidity, good operability | 35 | 43 | 0.027 |
| B: Slightly soft or slightly hard, general operation | 10 | 2 |  |
| **Knob operation** |  |  |  |
| A: Flexible | 42 | 45 | 0.242 |
| B: Fair, with certain resistance | 3 | 0 |  |
| **Sharp angle adaptability** |  |  |  |
| A: Good and easy to pass the tip of the scope | 41 | 45 | 0.117 |
| B: Fair and the tip of the scope can narrowly pass | 4 | 0 |  |
| **4.2.2 Auxiliary features** |  |  |  |
| **Air supply** |  |  |  |
| A: Sensitive, with moderate air supply | 43 | 45 | 0.494 |
| B: Relatively sensitive, with more or less air supply | 2 | 0 |  |
| **Water supply** |  |  |  |
| A: Sensitive and can effectively clean the scope | 41 | 45 | 0.117 |
| B: Relatively sensitive and can clean the scope | 4 | 0 |  |
| **Suction** |  |  |  |
| A: Sensitive, with a moderate suction volume | 44 | 45 | 1.000 |
| B: Relatively sensitive, with more or less suction volume | 1 | 0 |  |

##### 2.3 Efficiency of observation and potential EMR procedure

The total operating time, insertion time and withdrawal time were longer in the disposable group and the differences were statistically significant (*P*<0.01). The polyp(s) detection rates were 17/45 (37.8%) in experimental group and 20/45 (44.4%) in control group, whereas no significant difference was showed. 13 subjects in the experimental group and 17 subjects in the control group detected polyps were required to EMR procedure due to the relatively larger diameter (5mm-15mm). There was no significant difference about the age of those conducted EMR. The total number of polyps via EMR were 26 in these 13 subjects and 23 in the other group. As for the pathologic results, the fundic gland polyp occupied the majority, followed by Hyperplastic polyp. There were 1 low grade intraepithelial neoplasia (LGIN) case in the experimental group and 2 cases in the control group. No significant difference of the proportion of pathologic results. The En-bloc resection rate was the same 100% in both groups (26/26 vs 23/23, 95%CI: 0.8713,1.0000). Nevertheless, the procedure time of EMR for each polyp was significantly longer in the disposable group (*P*<0.01). **(Table 5)**

**Table 5.**
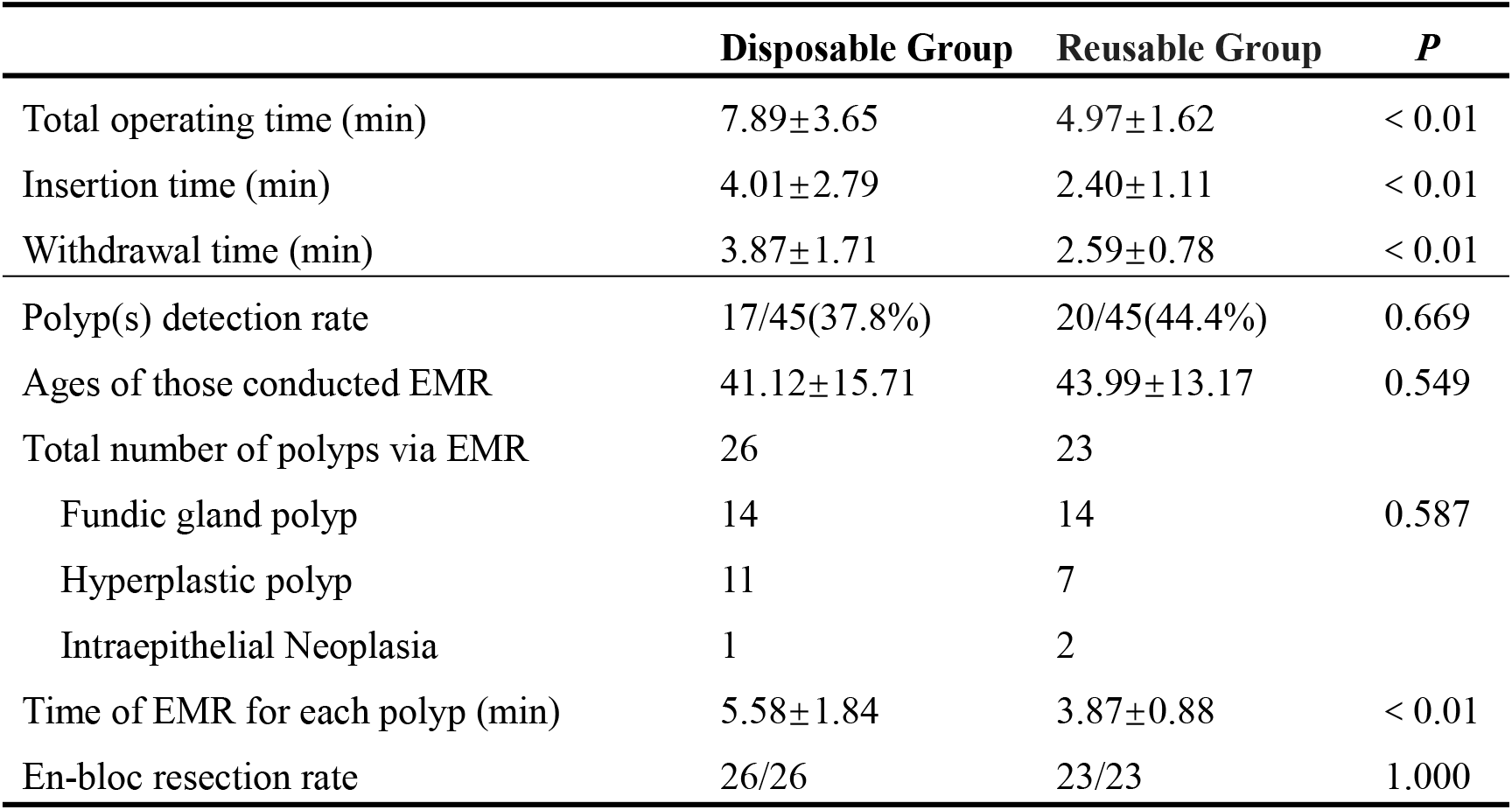
Comparison of the efficiency of observation and potential EMR procedure.

##### 2.4 Safety evaluation

No intraoperative bleeding, delayed bleeding, perforation was showed in both groups (0/45; 95% CI: 0.0000, 0.0787). The device failure/malfunction rate and the incidence of any other adverse events during the study period were 0.0% (0/45; 95% CI: 0.0000, 0.0787) in both groups. Before, during, 10 ± 5 mins and 1 h ± 15 mins after the procedure, no differences were statistically significant between two groups about stability evaluation (Diastolic blood pressure, Systolic blood pressure and heart rate). **(Table 6)**

**Table 6.** Comparison of the safety and stability evaluation between two groups.

|  | **Disposable Group** | **Reusable Group** | ***P*** |
| --- | --- | --- | --- |
| Intraoperative bleeding | 0/45 | 0/45 | NS |
| Delayed bleeding | 0/45 | 0/45 | NS |
| Perforation | 0/45 | 0/45 | NS |
| Equipment malfunction/defect | 0/45 | 0/45 | NS |
| Other adverse events | 0/45 | 0/45 | NS |
| **Stability evaluation** |  |  |  |
| **Diastolic blood pressure** |  |  |  |
| Before procedure | 42/45 | 44/45 | 0.616 |
| During procedure | 40/45 | 44/45 | 0.205 |
| 10 ± 5 mins after procedure | 43/45 | 44/45 | 1.000 |
| 1 h ± 15 mins after procedure | 44/45 | 45/45 | 1.000 |
| **Systolic blood pressure** |  |  |  |
| Before procedure | 41/45 | 43/45 | 0.677 |
| During procedure | 40/45 | 43/45 | 0.434 |
| 10 ± 5 mins after procedure | 43/45 | 44/45 | 1.000 |
| 1 h ± 15 mins after procedure | 44/45 | 45/45 | 1.000 |
| **Heart rate** |  |  |  |
| Before procedure | 36/45 | 41/45 | 0.230 |
| During procedure | 34/45 | 40/45 | 0.167 |
| 10 ± 5 mins after procedure | 40/45 | 42/45 | 0.714 |
| 1 h ± 15 mins after procedure | 43/45 | 45/45 | 0.494 |

## Discussion

The number of gastrointestinal endoscopic procedures performed in China annually is estimated to be 60 million. Moreover, endoscopes incur high capital costs in the form of purchase, repair, reprocessing, storage and disinfection, which often lead to be prohibitive. Regarding to the volumes for diagnosis and treatment via endoscopes home and abroad, single-use endoscopes might be a cost-effective alternative to that of conventional endoscopes. Nevertheless, comparative technical efficiency and safety between the two types have not been widely investigated in gastrointestinal societies^9^.

This non-randomized, controlled, open-label and noninferiority clinical trial is the first to identify and evaluate the efficiency, efficacy and safety of gastroscopy examination and potential endoscopic mucosal rection (EMR) procedure. The success rate of acceptable image quality via photographing iconic anatomical sites between two groups was 100.0% and the lower limit of the 95%CI (-7.8654%, 7.8654%) was larger than the noninferiority margin of -8%. Despite there were slightly lower grades between two groups in the index of image acquisition, image quality, brightness, contrast and sharpness, the image quality met basic clinical demands, especially considering the grades of A (good) and B (relatively good) were both qualified. Our study suggested that the image quality in the disposable group was noninferior to that in the reusable group.

As to accepted endoscopic maneuverability, the flexibility (knob operation, and sharp angle adaptability) rate was 100% in both groups, which was considering noninferior. Yet the control group had a better rating in terms of endoscopy body rigidity. Batches of these novel disposable endoscopes materials, familiarity with the traditional gastroscopes and the subjective operation experience of the endoscopists might lead to the differences. The acceptable auxiliary feature (air supply, water supply, and suction) rate was 100% in both groups, suggesting good and noninferior auxiliary performance. Consequently, the eligible scope cleaning (self-cleaning) and liquid or food residue removal from the site (site-cleaning) in these disposable devices might ensure successful examination or treatment.

The operating time consuming is also a valuable and important indicator to indirectly reflect the comprehensive quality of the gastroscope. The total operating time, insertion time, and withdrawal time were all dramatically shorter in the reusable group than that in the disposable group. Proficiency and familiarity with the conventional reusable gastroscopes among the well-trained and experienced endoscopists might be prone to these results. Additionally, when the investigators were operating these novel devices, more time would be consumed to observe and evaluate the performance at the beginning. As the endoscopists became assured of the feasibility of conducting the single-use gastroscope, decreased time consuming in mid-late period of the study could further confirm. Our results consist with the published study^3^.

The polyp(s) detection rate (PDR) was not statistically different between two groups ensured high-level examining quality and avoided missing diagnosis to some extent. 13 subjects in the experimental group and 17 subjects in the control group detected polyps were required to EMR procedure due to the relatively larger diameter (5mm-15mm). ESGE recommends that the goals of endoscopic mucosal resection (EMR) are to achieve a completely snare-resected lesion in the safest minimum number of pieces, with adequate margins and without need for adjunctive ablative techniques^10^. In order to minimize the selective bias in the population, subgroup analysis was conducted. No between-group significant diversity was founded in terms of the ages of those who conducted EMR procedures. The procedure time of EMR for each polyp was significantly longer in disposable group comparing with the reusable group. No previous studies related to disposable endoscopes in EMR, also the efficacy and safety of these single-use devices in EMR procedures remained unclear. Hence, it was quite possible that even the very experienced endoscopic experts would conduct the inventions much more slowly and cautiously, which also leads to an increase in time-consuming. What makes us feel optimistic is that the En-bloc resection rate was the same 100% in both groups (26/26 vs 23/23, 95%CI: 0.8713,1.0000), suggesting that the efficacy related to EMR procedure in the experimental group was equivalent to that in the control group.

This study showed no intraoperative bleeding, delayed bleeding, perforation or other study-related adverse events among 90 patients, indicating disposable gastroscopy might not be immediately or delayed harmful in both groups and indirectly proving the safety. No dramatic fluctuations in blood pressure or heart rate were showed in perioperative period, indicating that gastroscopy was well-tolerated in both groups and demonstrating good operational stability of the gastroscopes used in this study.

The widespread application of gastrointestinal endoscopy in diagnostic and therapeutic the disorders is accompanied by huge expense in purchasing, maintenance, reprocessing and disinfection. At present, the digestive tract endoscope products in the market all over the world have many functions, small size, many and slender internal pipelines, complex product structure, and many kinds of cavities and semi closed cavities. Reprocessing of conventional endoscope typically consists of 8 steps: precleaning, leak testing, manual cleaning, irrigation after cleaning, visual examination, advanced disinfection, rinsing after high-level disinfection, and drying^11^, yet residual droplets, microbial growth and biofilm formation might still exist in gastrointestinal endoscopes after reprocessing and drying. During immersion, the disinfectant at some positions may not be fully released, which is likely to lead to incomplete disinfection. Regarding to the characteristics of these disposable endoscopic devices, an alternative option might be given to overcome these disadvantages and provide non-inferior performance.

Conventional reusable endoscopic equipment is high-precision instrument that requires careful maintenance. Not only routine endoscopy, but also various therapeutic operations may cause loss of components. Previous studies suggested eligible performance of disposable gastroscope in routine examination comparing the reusable gastroscope^3^. In this study, progressive potential EMR operations were conducted and analyzed. Given our findings, we suggested that the single-use gastroscopic devices were also non-inferior to traditional ones in examination and EMR treatment, which can be applied for some special populations or those who demand one-time endoscopy for personal reasons.

In addition, the disposable gastroscopes are playing an important role in some special circumstances when considering the potential contamination and high expense. For those with sever immune deficiency diseases or low immunity condition, disposable endoscopy is sterile and may provide an appropriate resolution to minimize possible transmission or cross infection. Disposable gastroscopes are also an acceptable bedside device in the wards, the Emergency area or Intensive Care Unit when considering the emergency situations. Furthermore, it was widely known that the COVID-19 epidemic abruptly restricted gastrointestinal endoscopy services during the first wave of pandemic. The volume of endoscopies was markedly reduced because it was strongly recommended that the nonurgent endoscopies should be postponed or canceled to prevent the spread of SARS-CoV-2 by academic societies on endoscopy, and the reduction was also due to decreased scale of outpatients and cancellations by patients, indicating that individuals were highly aware of the risk of SARS-CoV-2 cross infection by endoscopy^12^. Published study^13^ suggested that the application of a disposable endoscope was also a preferred option for minimizing the risk of contact transmission during the severe and wide-spread epidemic in 2020. As a consequence, we consider that disposable endoscopes could also be applied in battlefield, mobile clinic or special infectious wards even in the pandemic.

There are some limitations of note. Firstly, this is a non-randomized controlled study. We fully regarded the individual wishes among these enrolled patients and written inform contents were completely explained and obtained, whereas selection bias is inevitable to some content comparing with RCTs. Therefore, we restricted the criteria in the design process especially some factors which were seemed to be potential confounders. We conducted an advanced subgroup analysis to minimize the differences on the baseline between two groups. The blinding method was also used to control the bias. We arranged the investigators who were not directly involved in clinical decision-making to complete the Case Report Form (CRF) and the statistical analysis. Secondly, this is a non-inferiority trial. The total performance of the disposable devices was not quite as good as that of the conventional gastroscope in mature GI endoscopy society. We believe that a much better user experience could be provided in the future with the developments and improvements in the field of image quality and maneuverability. The cost-effectiveness studies should be conducted and the expense would decrease over time as production scales up^3^. However, the plastic waste might be generated if the devices are not recycled. Some organizations are concerned that the widespread promotion and application of those disposable devices might implicate the environment^14^. How to weight the benefits and the risks? This question could be addressed from four perspectives—clinical, financial, innovation and environmental protection.

## Conclusion

In summary, given the overall efficiency and safety profile, non-inferior technical performance of observation and endoscopic mucosal resection operation, the disposable gastroscopes might represent an alternative to conventional reusable gastroscopes and this innovation might be a significant advancement in the field of gastrointestinal endoscopy.

## Data Availability

All data produced in the present work are contained in the manuscript

